# Pectoralis muscle dosimetry and post-treatment rehabilitation utilization for early-stage breast cancer patients

**DOI:** 10.1101/2023.04.28.23289275

**Authors:** Jamie S.K. Takayesu, Shannon J. Jiang, Robin Marsh, Alexander Moncion, Sean R. Smith, Lori J. Pierce, Reshma Jagsi, David B. Lipps

## Abstract

Up to 50% of women treated for localized breast cancer will experience some degree of arm or shoulder morbidity. While radiation is thought to contribute to this morbidity, the mechanism remains unclear. Prior studies have shown biologic and radiographic changes in the pectoralis muscles after radiation. This study thus aims to investigate the relationship between radiation to the pectoralis muscles and referrals for rehabilitation services post-treatment for arm and shoulder morbidity. A retrospective 1:1 matched case-control study was conducted for breast cancer patients who were and were not referred for breast or shoulder rehabilitation services between 2014-2019 at a single academic institution. Patients were included if they had a lumpectomy and adjuvant radiation. Patients who underwent an axillary lymph node dissection (ALND) were excluded. Cohorts were matched based on age, axillary surgery, and use of radiation boost. Muscle doses were converted to EQD2 assuming an α/β ratio of 2.5 and were compared between the two groups. In our cohort of 50 patients of a median age 60 years (interquartile range (IQR) 53-68 years), 36 patients (72%) underwent a sentinel lymph node biopsy (SLNB) in addition to a lumpectomy. While pectoralis muscle doses were generally higher in those receiving rehabilitation services, this was not statistically significant. Pectoralis major V20-40Gy reached borderline significance, as did pectoralis major mean dose (17.69Gy vs. 20.89Gy, p=0.06). In this limited cohort of patients, we could not definitively conclude a relationship between pectoralis muscle doses and use of rehabilitation services. Given the borderline significant findings, this should be further investigated in a larger cohort.

## Introduction

Breast cancer is the most common cancer in women with an incidence rate of 129.7 per 100,000 women per year.^1^ Prognosis continues to improve over time,^2^ which is why survivorship is becoming ever more critical. One important component of survivorship is the identification and management of shoulder or arm morbidity, which affects up to 50% of women treated for breast cancer^3,4^ and is associated with poorer quality of life.^5^ Shoulder morbidity includes pain, restricted range of motion, stiffness, fibrosis, lymphedema and axillary web syndrome.^3,5–7^ While shoulder morbidity is less severe after breast conserving therapy (BCT), which consists of breast conserving surgery and radiation therapy, compared to mastectomy,^8^ rates are still as high as 30-50%.^3,6^

The mechanism behind shoulder morbidity after radiation is still unclear. Key shoulder muscles like the pectoralis major, pectoralis minor, latissimus dorsi and teres major receive the highest doses of radiation with 3D conformal radiation therapy (3DCRT) for whole breast radiation.^9^ For the pectoralis muscles specifically in patients treated with radiation, there are biologic and radiographic findings suggestive of radiation-induced changes. These changes include increased inflammation and muscular atrophy,^10^ increased muscle stiffness^11,12^ and decreased pectoralis muscle volume.^13,14^ While these prior studies observed changes to the pectoralis muscles after radiation, it is still unknown whether these findings translate into clinically meaningful endpoints.

The primary management for shoulder morbidity following surgery or radiation is with cancer rehabilitation services. These services significantly improve outcomes after breast cancer treatment, particularly when patients are referred early.^8^ Patients with clinically perceptible arm and shoulder morbidity are often referred by their oncologic team to a physiatrist or other rehabilitation provider for assessment and treatment. Aside from clinical judgment, there are few tools to aid in the early detection of arm and shoulder morbidity. One readily available potential clinical decision aid that could assist in patient referral to rehabilitation services is radiation dosimetric data, particularly that of the pectoralis muscles, if a correlation between radiation dose delivered and shoulder morbidity was known.

The relationship between pectoralis muscle radiation dose and subsequent shoulder morbidity requiring physiatrist assessment and intervention has yet to be explored after radiation therapy. Therefore, the aim of this study was to investigate the relationship between radiation dose delivered to the pectoralis major and pectoralis minor and referrals for cancer rehabilitation services post-treatment.

## Materials and Methods

Institutional review board approval was obtained for this retrospective study (HUM00202558). We identified a 1:1 matched case-control cohort of females with breast cancer treated with BCT who were or were not referred for breast or shoulder rehabilitation services between 2014-2019 at a single academic institution. Cohorts were matched based on age (within 5 years), axillary surgery (none vs. SLNB), and use of radiation boost. Patients were included if they had a lumpectomy with or without a SLNB and adjuvant whole breast irradiation (WBI), and a minimum of 3 years of follow-up with providers within our medical system. Patients were excluded if they had a mastectomy, ALND, reconstructive surgery on the ipsilateral breast, or a recurrence within 3 years of completing breast radiation. Patients were also excluded if radiation plans were not accessible or if they were undergoing rehabilitation for the ipsilateral arm or shoulder prior to starting radiation. Clinical data were retrospectively extracted from the electronic medical record (EMR).

### Radiation therapy

After initial consultation, all patients underwent a CT simulation in the supine position. Radiation plans were retrospectively reviewed in Aria Eclipse version 16.00.00 (Varian Medical Systems, Palo Alto, CA). All plans were created using a 3DCRT technique. Radiation boosts were delivered either using photons or electrons. Two investigators independently (J.T. and S.J.) contoured the pectoralis major and pectoralis minor muscles as detailed in Lipps et al.^9^ All contours were verified by a single investigator (J.T.) for quality assurance. Prespecified dose-volume metrics were extracted from the planning software. All doses were converted to EQD2 assuming an α/β ratio of 2.5.

### Rehabilitation services

The EMR was reviewed to identify patients who were referred for rehabilitation services at least one month after completing WBI. Rehabilitation services included physical therapy (PT), occupational therapy (OT) or Physical Medicine and Rehabilitation (PMR).

Patients who underwent rehabilitation for the ipsilateral arm or shoulder prior to starting radiation therapy were excluded from this study. Diagnosis codes and free text on the referral and rehabilitation notes were used to identify whether a patient was referred for a problem reasonably relating to sequelae of BCT. Referral free-text and rehabilitation notes were reviewed for the following signs and symptoms: lymphedema, shoulder pain, arm pain, restricted shoulder or arm range of motion, breast pain, axillary webbing, and scar tightness.

Those referred for scar tightness had to have an additional reason for rehabilitation referral to be considered part of the rehabilitation group. Receiving rehabilitation services for reasons outside of the ipsilateral breast, arm or shoulder did not count toward distinguishing between the rehabilitation versus no rehabilitation cohort.

### Statistics

A Fisher’s exact test or Pearson’s Chi-squared test were used to evaluate differences in categorical variables between groups. A Wilcoxon rank sum test was used to determine differences in continuous variables, including whether pectoralis major or pectoralis minor doses (mean, V50Gy, V45Gy, V40Gy, V35Gy, V30Gy, V20 Gy) were different between the two groups. A linear regression model was used to conduct all univariate analyses. All p-values are two-sided and were considered statistically significant if less than 0.05. All statistical analyses were performed using R version 4.2.2 (R Foundation for Statistical Computing, Vienna, Austria).

## Results

### Patient characteristics

Baseline characteristics of the 50 patients evaluated in our study are detailed in **Table 1**. In our cohort, the median age was 60 years (interquartile range [IQR] 54-67 years), and most patients were White (90%). 29 patients (58%) had never smoked, and the median BMI was 30kg/m^2^ (IQR 27-36 kg/m^2^). Of the sample, 36 patients (72%) underwent SLNB in addition to lumpectomy (**Table 2**). Median tumor size was 1cm (IQR 1-2cm). Patients had Stage 0 (n=14, 28%), I (n=24, 48%), II (n=11, 22%), or III (n=1, 2%) breast cancer.

**Table 1.**
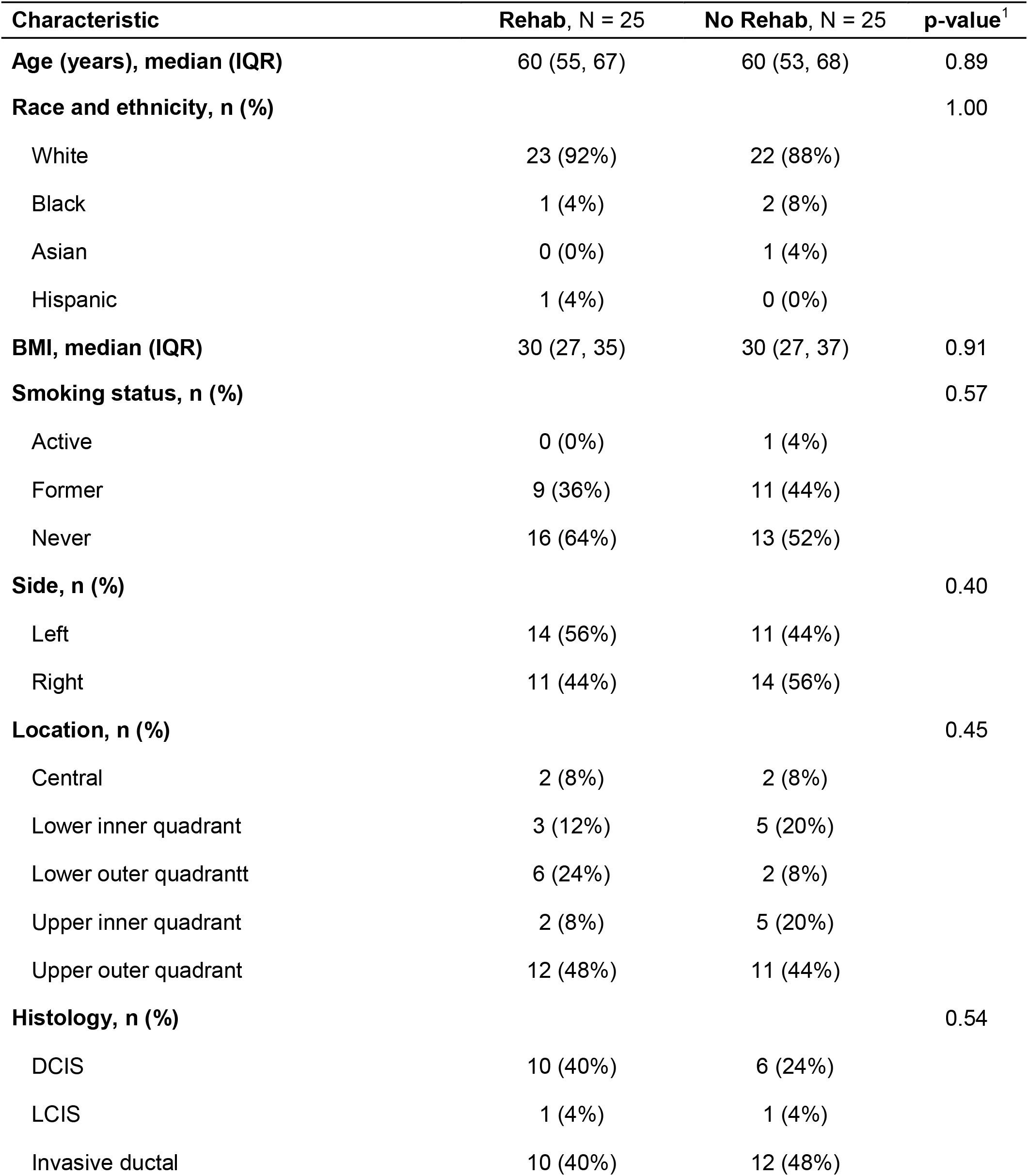
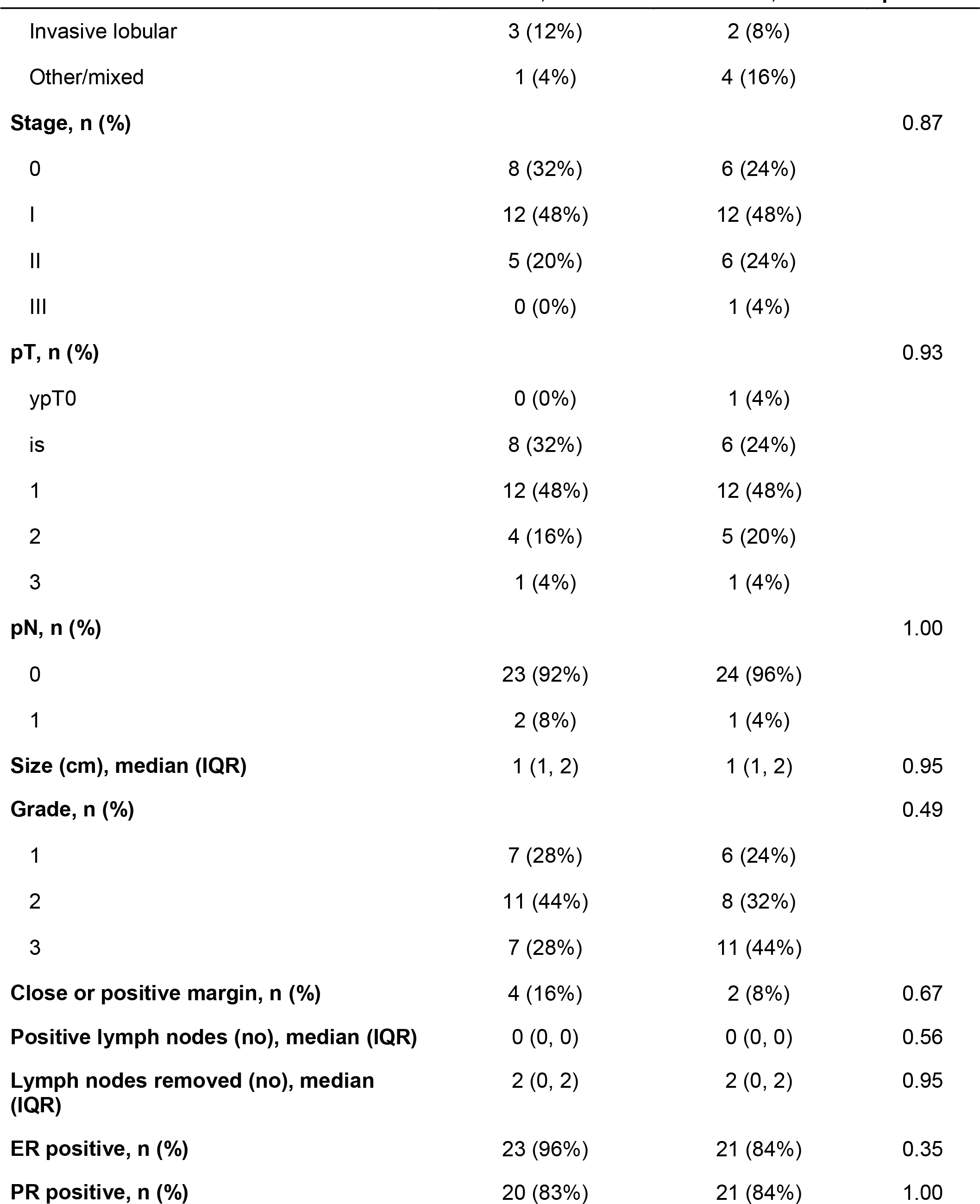
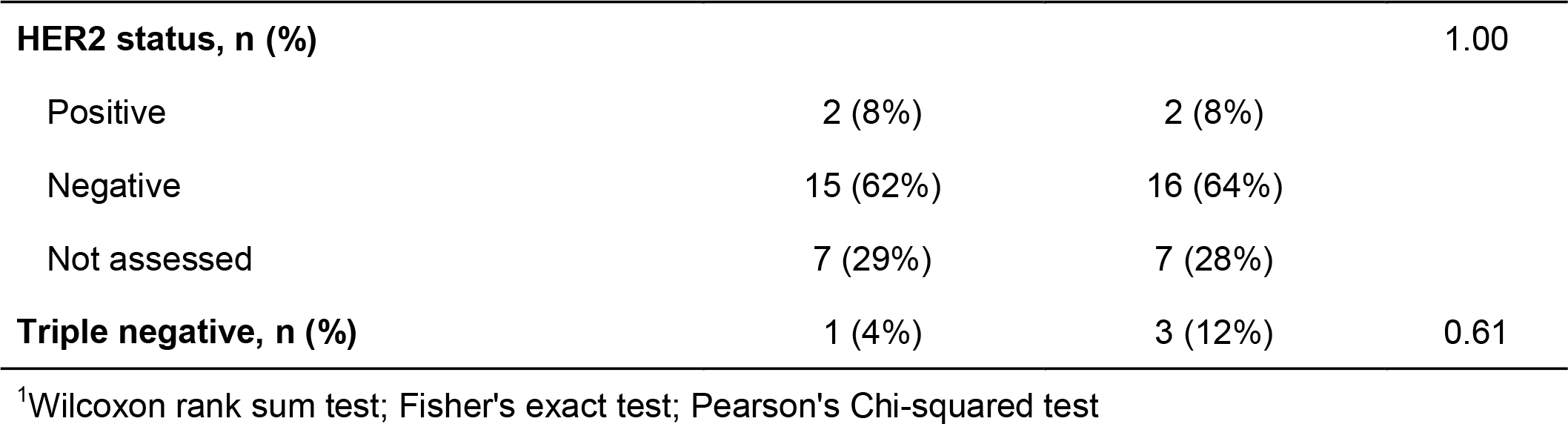
Baseline characteristics

| Characteristic | Rehab, N = 25 | No Rehab, N = 25 | p-value<sup>1</sup> |
| --- | --- | --- | --- |
| **Age (years), median (IQR)** | 60 (55, 67) | 60 (53, 68) | 0.89 |
| **Race and ethnicity, n (%)** |  |  | 1.00 |
| White | 23 (92%) | 22 (88%) |  |
| Black | 1 (4%) | 2 (8%) |  |
| Asian | 0 (0%) | 1 (4%) |  |
| Hispanic | 1 (4%) | 0 (0%) |  |
| **BMI, median (IQR)** | 30 (27, 35) | 30 (27, 37) | 0.91 |
| **Smoking status, n (%)** |  |  | 0.57 |
| Active | 0 (0%) | 1 (4%) |  |
| Former | 9 (36%) | 11 (44%) |  |
| Never | 16 (64%) | 13 (52%) |  |
| **Side, n (%)** |  |  | 0.40 |
| Left | 14 (56%) | 11 (44%) |  |
| Right | 11 (44%) | 14 (56%) |  |
| **Location, n (%)** |  |  | 0.45 |
| Central | 2 (8%) | 2 (8%) |  |
| Lower inner quadrant | 3 (12%) | 5 (20%) |  |
| Lower outer quadrant | 6 (24%) | 2 (8%) |  |
| Upper inner quadrant | 2 (8%) | 5 (20%) |  |
| Upper outer quadrant | 12 (48%) | 11 (44%) |  |
| **Histology, n (%)** |  |  | 0.54 |
| DCIS | 10 (40%) | 6 (24%) |  |
| LCIS | 1 (4%) | 1 (4%) |  |
| Invasive ductal | 10 (40%) | 12 (48%) |  |
| Invasive lobular | 3 (12%) | 2 (8%) |  |
| Other/mixed | 1 (4%) | 4 (16%) |  |
| **Stage, n (%)** |  |  | 0.87 |
| 0 | 8 (32%) | 6 (24%) |  |
| I | 12 (48%) | 12 (48%) |  |
| II | 5 (20%) | 6 (24%) |  |
| III | 0 (0%) | 1 (4%) |  |
| **pT, n (%)** |  |  | 0.93 |
| ypT0 | 0 (0%) | 1 (4%) |  |
| is | 8 (32%) | 6 (24%) |  |
| 1 | 12 (48%) | 12 (48%) |  |
| 2 | 4 (16%) | 5 (20%) |  |
| 3 | 1 (4%) | 1 (4%) |  |
| **pN, n (%)** |  |  | 1.00 |
| 0 | 23 (92%) | 24 (96%) |  |
| 1 | 2 (8%) | 1 (4%) |  |
| **Size (cm), median (IQR)** | 1 (1, 2) | 1 (1, 2) | 0.95 |
| **Grade, n (%)** |  |  | 0.49 |
| 1 | 7 (28%) | 6 (24%) |  |
| 2 | 11 (44%) | 8 (32%) |  |
| 3 | 7 (28%) | 11 (44%) |  |
| **Close or positive margin, n (%)** | 4 (16%) | 2 (8%) | 0.67 |
| **Positive lymph nodes (no), median (IQR)** | 0 (0, 0) | 0 (0, 0) | 0.56 |
| **Lymph nodes removed (no), median (IQR)** | 2 (0, 2) | 2 (0, 2) | 0.95 |
| **ER positive, n (%)** | 23 (96%) | 21 (84%) | 0.35 |
| **PR positive, n (%)** | 20 (83%) | 21 (84%) | 1.00 |
| **HER2 status, n (%)** |  |  | 1.00 |
| Positive | 2 (8%) | 2 (8%) |  |
| Negative | 15 (62%) | 16 (64%) |  |
| Not assessed | 7 (29%) | 7 (28%) |  |
| **Triple negative, n (%)** | 1 (4%) | 3 (12%) | 0.61 |

<sup>1</sup>Wilcoxon rank sum test; Fisher's exact test; Pearson's Chi-squared test

**Table 2.** Treatment details

| **Characteristic** | **Rehab, N = 25** | **No Rehab, N = 25** | **p-value<sup>1</sup>** |
| --- | --- | --- | --- |
| **Surgery, n (%)** |  |  |  |
| Lumpectomy | 25 (100%) | 25 (100%) |  |
| **Axillary Surgery, n (%)** |  |  | 1.00 |
| None | 7 (28%) | 7 (28%) |  |
| Sentinel node | 18 (72%) | 18 (72%) |  |
| **Neoadjuvant chemo, n (%)** | 0 (0%) | 2 (8%) | 0.49 |
| **Endocrine therapy, n (%)** |  |  | 0.10 |
| Tamoxifen | 8 (32%) | 3 (12%) |  |
| Aromatase Inhibitor | 12 (48%) | 11 (44%) |  |
| None | 5 (20%) | 11 (44%) |  |
| **Adjuvant chemo, n (%)** | 4 (16%) | 4 (16%) | 1.00 |
| **Breast dose (Gy), median (IQR)** | 43 (43, 50) | 43 (43, 50) | 0.85 |
| **Boost dose (Gy), median (IQR)** | 10 (2, 10) | 10 (10, 10) | 0.98 |
| **Fractionation, n (%)** |  |  | 0.78 |
| Conventionally fractionated | 12 (48%) | 11 (44%) |  |
| Hypofractionated | 13 (52%) | 14 (56%) |  |
| **Duration of radiation (days), median (IQR)** | 29 (26, 40) | 29 (26, 40) | 1.00 |
| **Radiation Fields, n (%)** |  |  | 1.00 |
| Tangents | 18 (72%) | 18 (72%) |  |
| High Tangents | 6 (24%) | 6 (24%) |  |
| Regional nodal irradiation | 1 (4%) | 1 (4%) |  |

<sup>1</sup> Pearson's Chi-squared test; Fisher's exact test; Wilcoxon rank sum test

### Radiation

A representative plan with pectoralis muscle contours is shown in **Figure 1**. Radiation was either moderately hypofractionated (n=27, 54%) or conventionally fractionated (n=23, 46%) (**Table 2**). A typical moderately hypofractionated plan was 42.56Gy in 16 fractions, and a typical conventionally fractionated plan was 50Gy in 25 fractions. Only one patient in each group received regional nodal irradiation (RNI), while 6 patients in each group received a boost.

**Figure 1.**
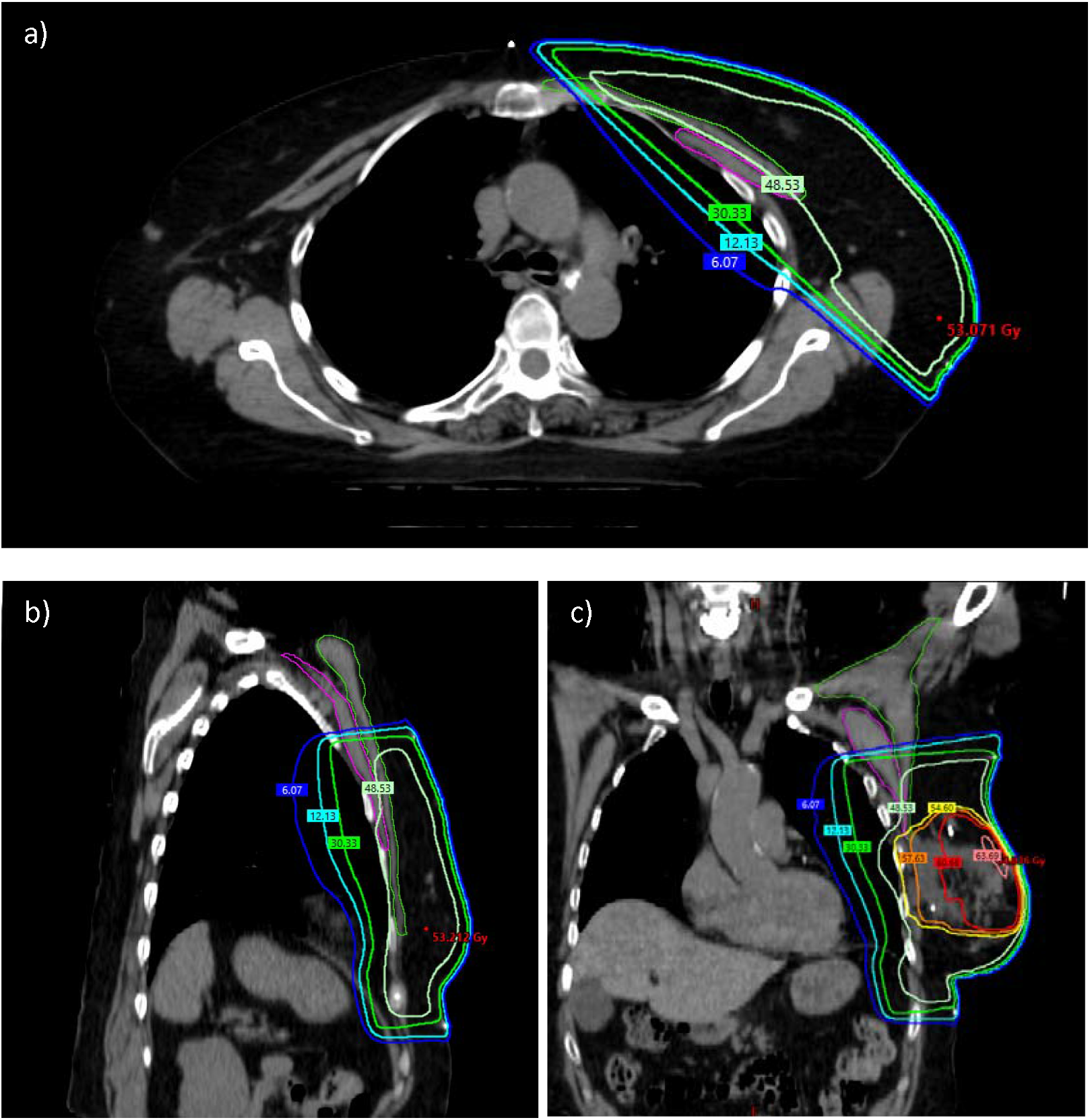
Representative conventionally fractionated radiation plan with a photon boost. Representative axial, sagittal and coronal slices are shown in panels a, b, and c, respectively. The pectoralis major muscle is outlined in green, and the pectoralis minor muscle is outlined in magenta. Isodose lines are as labeled.

### Rehabilitation services

The median time from end of WBI to rehabilitation referral was 4 months (IQR 3-9 months). Nine women (38%) were referred for rehabilitation within 6 months of completing radiation, 21 women (84%) were referred within 12 months, and 4 women (16%) were referred more than 12 months, with one woman being referred 44 months post-radiation. Most were referred to occupational therapy (n=22, 88%), and few were referred directly to PMR (n=3, 12%); thus 25 women received shoulder rehabilitation services. The most common primary rehabilitation diagnoses were lymphedema (n=12, 48%), restricted shoulder range of motion (n=2, 8%), breast pain (n=3, 12%), shoulder pain (n=4, 16%), and scar management (n=4, 16%). Ten women (40%) were assessed or treated for shoulder morbidity, which was defined as restricted shoulder range of motion, breast or chest wall pain or shoulder pain.

No patient or tumor characteristics were associated with increased likelihood of a rehabilitation referral on univariate analysis (**Supplementary Table 1**). However, univariate analysis of treatment characteristics showed that patients who did not receive endocrine therapy were less likely to be referred for rehabilitation services compared to those who received tamoxifen (OR 0.17, 95% confidence interval [CI] 0.03-0.86, p=0.041).

### Dose-volume metrics and rehabilitation referrals

There was a trend of higher doses to the pectoralis major in those referred for rehabilitation, but none were statistically significant between the two groups (**Table 3, Figure 2**). Pectoralis major mean dose (median 20.89Gy vs. 17.69Gy, p=0.06), V40Gy (37.80% vs. 32.69%, p=0.09), V35Gy (38.76% vs. 34.43%, p=0.09), and V20Gy (41.05% vs. 36.82%, p=0.06) were higher in the rehabilitation group compared to the no rehabilitation group, respectively. However, the comparisons were not statistically different.

**Table 3.** Pre-specified dose-volume metrics compared between the rehabilitation (PT) and no rehabilitation (No PT) groups. Pmajor = pectoralis major, Pminor = pectoralis minor.

| **Dose-Volume Metric** | **Rehab, N = 25<sup>1</sup>** | **No Rehab, N = 25<sup>1</sup>** | **p-value<sup>2</sup>** |
| --- | --- | --- | --- |
| **Breast V95% (%)** | 94.10 (91.02, 95.37) | 92.19 (84.62, 94.97) | 0.21 |
| **Heart mean dose (Gy)** | 0.55 (0.32, 0.66) | 0.40 (0.26, 0.66) | 0.32 |
| **PMajor Dmean (Gy)** | 20.89 (18.92, 23.63) | 17.69 (17.19, 20.69) | 0.06 |
| **PMajor Dmax (Gy)** | 59.66 (52.53, 62.26) | 60.29 (57.44, 63.11) | 0.58 |
| **PMajor D0.1cc (Gy)** | 58.98 (52.27, 61.80) | 60.10 (57.16, 62.60) | 0.50 |
| **PMajor V55Gy (%)** | 1.65 (0.00, 7.48) | 4.07 (0.89, 6.39) | 0.35 |
| **PMajor V50Gy (%)** | 12.32 (5.84, 14.58) | 10.29 (5.27, 18.88) | 0.96 |
| **PMajor V45Gy (%)** | 33.79 (24.98, 37.23) | 28.68 (21.22, 35.19) | 0.25 |
| **PMajor V40Gy (%)** | 37.80 (33.29, 41.93) | 32.69 (29.87, 38.40) | 0.09 |
| **PMajor V35Gy (%)** | 38.76 (35.21, 43.28) | 34.43 (31.38, 39.61) | 0.09 |
| **PMajor V30Gy (%)** | 39.51 (36.73, 44.05) | 35.36 (32.42, 40.24) | 0.07 |
| **PMajor V20Gy (%)** | 41.05 (39.38, 45.33) | 36.82 (34.14, 41.28) | 0.06 |
| **PMinor Dmean (Gy)** | 29.25 (25.39, 35.64) | 26.17 (22.32, 30.71) | 0.32 |
| **PMinor Dmax (Gy)** | 53.15 (48.65, 57.35) | 54.43 (47.85, 58.45) | 1.00 |
| **PMinor D0.1cc (Gy)** | 51.80 (48.17, 56.61) | 54.01 (47.05, 57.27) | 0.85 |
| **PMinor V55Gy (%)** | 0.00 (0.00, 2.22) | 0.00 (0.00, 2.87) | 1.00 |
| **PMinor V50Gy (%)** | 1.43 (0.00, 10.77) | 3.32 (0.00, 14.51) | 0.75 |
| **PMinor V45Gy (%)** | 34.68 (11.04, 51.21) | 28.49 (7.32, 39.72) | 0.37 |
| **PMinor V40Gy (%)** | 52.97 (45.40, 70.02) | 49.12 (38.71, 60.96) | 0.26 |
| **PMinor V35Gy (%)** | 59.42 (51.30, 75.96) | 52.91 (46.73, 63.75) | 0.34 |
| **PMinor V30Gy (%)** | 61.85 (52.52, 76.89) | 54.36 (48.72, 64.96) | 0.35 |
| **PMinor V20Gy (%)** | 63.99 (54.43, 78.32) | 56.18 (52.09, 66.68) | 0.38 |

<sup>1</sup>Median (IQR)
<sup>2</sup>Wilcoxon rank sum test; Wilcoxon rank sum exact test

**Figure 2.**
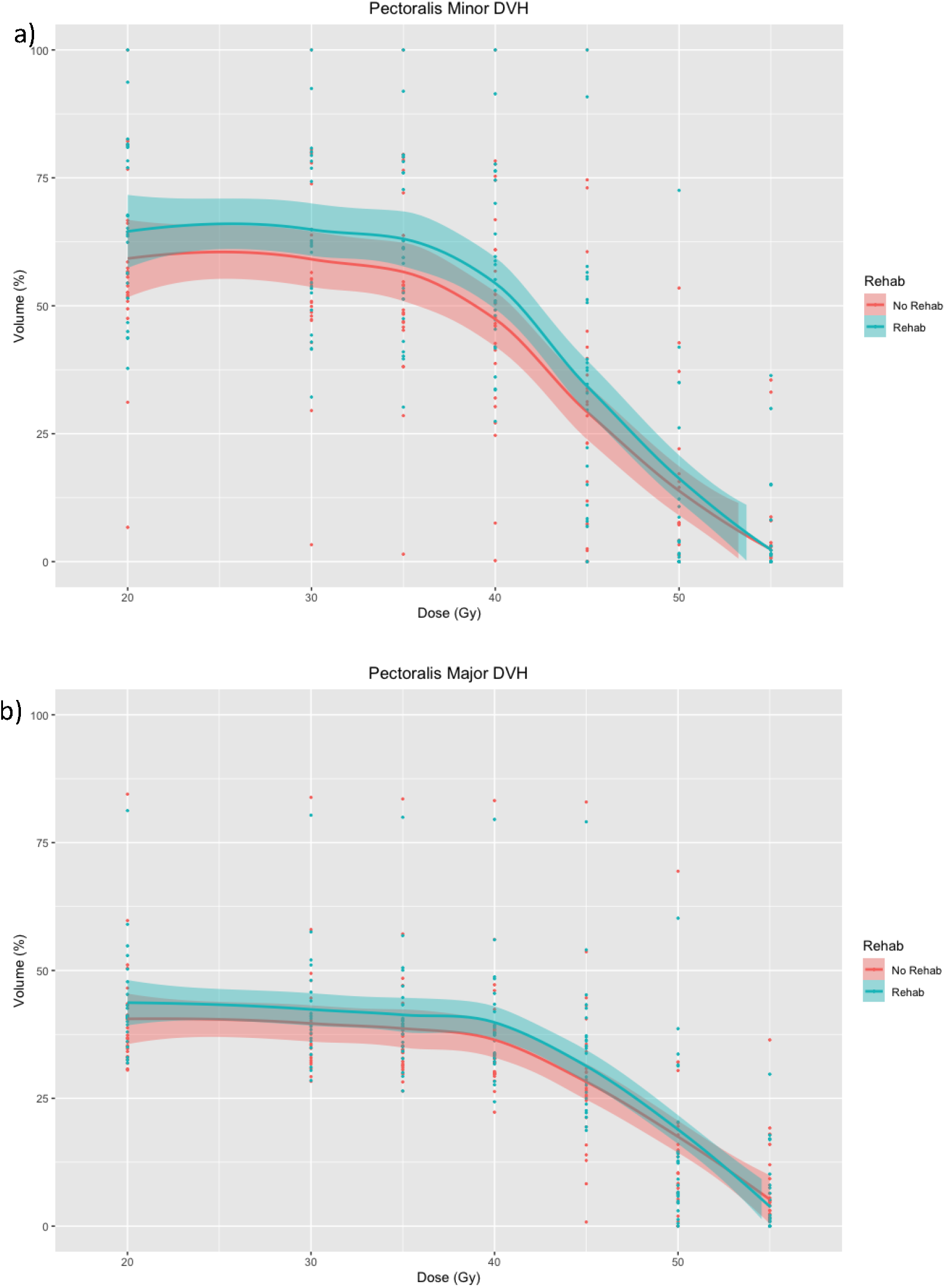
Summative dose-volume histogram for pectoralis minor (a) and pectoralis major (b). Shown in turquoise are those trend lines and 95% for the group who did and did not receive rehabilitation. Points represent individual patient data.

Similarly, there was no statistically significant difference in the pre-defined pectoralis minor dose-volume metrics between the two groups (**Table 3**).

## Discussion

Several studies have investigated the biologic and radiographic changes to the pectoralis muscles after radiation.^10–14^ To our knowledge, this was the first study attempting to correlate radiation doses to the pectoralis muscle with a clinically meaningful endpoint of requiring rehabilitation after radiation. We found that most women are referred for rehabilitation within one year of completing radiation therapy. While lymphedema was the most common reason women were referred for rehabilitation, shoulder morbidity was noted in 40% of the women who were referred for rehabilitation. We also found a trend toward higher pectoralis major mean dose and V20-40Gy in women referred for rehabilitation. While this finding was not statistically significant, it may warrant further investigation in a larger cohort of patients.

Around 40% of women will experience shoulder dysfunction after lumpectomy and radiation using modern techniques.^3^ Undergoing more extensive surgeries like a mastectomy or ALND result in worse shoulder function compared to less extensive surgeries like a lumpectomy or SLNB.^15^ It is poorly understood how radiation contributes to shoulder dysfunction. This has been indirectly addressed in large trials on omission of radiation including CALGB 9343, which randomized women over 70 years old to lumpectomy with or without WBI. Patient-reported arm or shoulder stiffness and breast pain were worse in those who received radiation compared to those who did not.^16^ EORTC 22922 and MA.20 investigated the utility of adding RNI to WBI, and neither trial detected a decrement to arm and shoulder stiffness by expanding radiation field.^17–19^

One important contributor to these changes in shoulder function and shoulder and breast pain could include changes in the pectoralis muscles after WBI. The pectoralis muscles were of particular interest in this cohort given that they receive moderate to high doses with reasonable heterogeneity between plans in this cohort of largely 2-field plans^9^. This raises concern for the involvement of the pectoralis muscles in shoulder dysfunction and pain, as breast radiation has previously been shown to cause inflammation, fibrosis and atrophy in the pectoralis muscles.^10–13^

Our research team previously showed that shear elastic modulus (a measure of muscle stiffness) was correlated to mean radiation dose to the pectoralis major^11^ and that an increase in shear elastic modulus between 30 days post-RT and 6 months post-RT was strongly correlated with mean RT dose to the pectoralis major.^12^ In this cohort of patients with early-stage breast cancer, pectoralis major and pectoralis minor doses were generally higher in those receiving rehabilitation services, though this was not statistically significant. Our study also demonstrated no difference in the use of rehabilitation services regardless of fractionation schema, which was similarly reported on in the START trials.^3^

When shoulder morbidity does occur, early exercise programs and rehabilitation services improve shoulder function, quality of life and cost effectiveness.^20–22^ The fact that only 38% of women who required rehabilitation services in our cohort were referred within 6 months post-radiation might signal that better tools are needed for early detection and referrals. At present, referrals to rehabilitation services for shoulder dysfunction are typically made by practitioners based on clinical symptoms. There are multiple decision aids to help clinicians with early detection of lymphedema,^23,24^ but there are few tools to measure early changes to shoulder stiffness, though ultrasound shear wave elastography appears promising.^11,12^ With limited decision support technology, additional clinical information is needed to identify patients with shoulder dysfunction, with the aim of earlier referrals to rehabilitation services. Pectoralis muscle dosimetry could feasibly be helpful in aiding these decisions proactively, particularly if it might identify patients without clear early clinical symptoms and who are at risk for developing shoulder pain and dysfunction later on.

Limitations of this study include a small sample size, though this was a cohort with detailed radiation planning, strict matching criteria, consistent muscle contouring and rehabilitation details. We intentionally excluded patients who had a mastectomy or ALND to avoid confounding surgical effects on rehabilitation needs. Further studies should be considered in these populations as well, as more invasive surgeries may have an additive effect on morbidity and post-mastectomy pain is a major source for referrals to PMR and PT; potential additive effects of radiation should be explored. The low number of patients with RNI is consistent with our referral and practice patterns. Including more patients with RNI could increase the variability in pectoralis doses, which could allow us to detect an association with rehabilitation services. Further studies are warranted in these settings.

The current study provides information on the patterns of referral and details on arm and shoulder morbidity after radiation at a large referral center. The study suggests a trend between pectoralis major dosimetry and shoulder and arm morbidity, however further study is needed. Continued efforts to clarify the pathophysiology of shoulder impairment after radiation could help with radiation planning considerations and earlier rehabilitation referrals, which could result in better quality of life for breast cancer survivors.

## Data Availability

All data produced in the present study are available upon reasonable request to the authors

---

**Supplementary Table 1.**
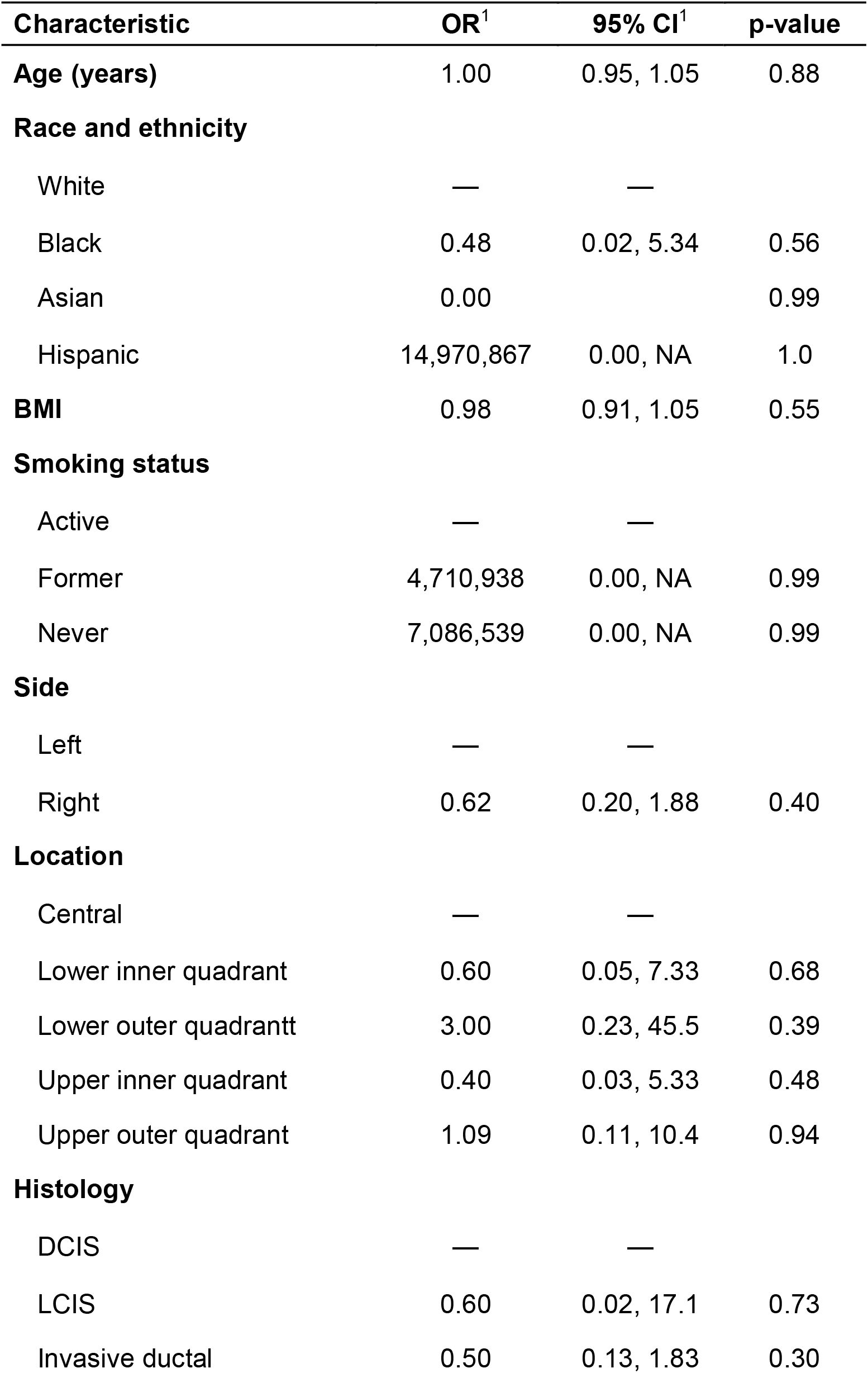
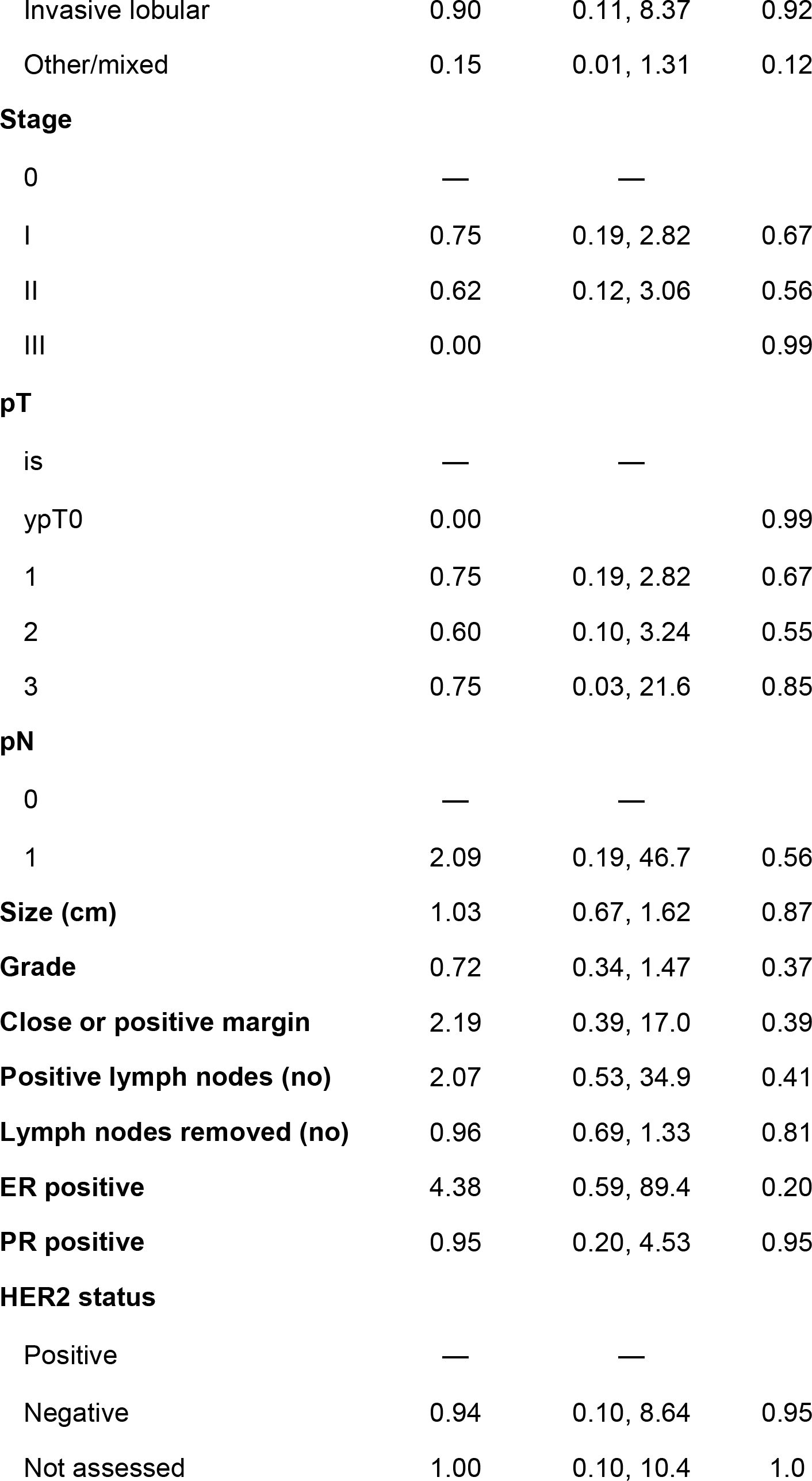
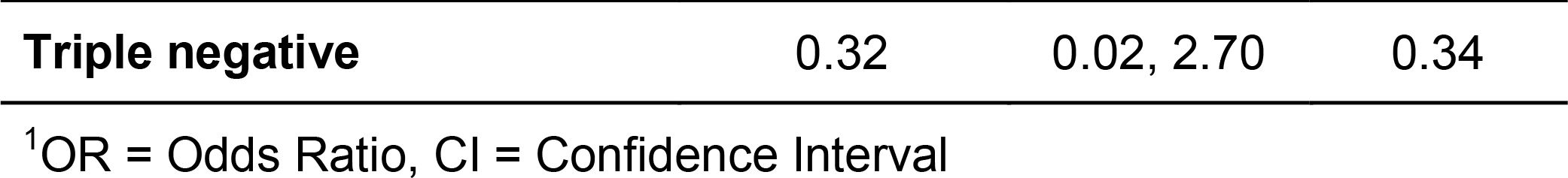
Univariate analysis of patient characteristics and use of rehabilitation services

| Characteristic | OR<sup>1</sup> | 95% CI<sup>1</sup> | p-value |
| --- | --- | --- | --- |
| **Age (years)** | 1.00 | 0.95, 1.05 | 0.88 |
| **Race and ethnicity** |  |  |  |
| White | — | — |  |
| Black | 0.48 | 0.02, 5.34 | 0.56 |
| Asian | 0.00 |  | 0.99 |
| Hispanic | 14,970,867 | 0.00, NA | 1.0 |
| **BMI** | 0.98 | 0.91, 1.05 | 0.55 |
| **Smoking status** |  |  |  |
| Active | — | — |  |
| Former | 4,710,938 | 0.00, NA | 0.99 |
| Never | 7,086,539 | 0.00, NA | 0.99 |
| **Side** |  |  |  |
| Left | — | — |  |
| Right | 0.62 | 0.20, 1.88 | 0.40 |
| **Location** |  |  |  |
| Central | — | — |  |
| Lower inner quadrant | 0.60 | 0.05, 7.33 | 0.68 |
| Lower outer quadrant | 3.00 | 0.23, 45.5 | 0.39 |
| Upper inner quadrant | 0.40 | 0.03, 5.33 | 0.48 |
| Upper outer quadrant | 1.09 | 0.11, 10.4 | 0.94 |
| **Histology** |  |  |  |
| DCIS | — | — |  |
| LCIS | 0.60 | 0.02, 17.1 | 0.73 |
| Invasive ductal | 0.50 | 0.13, 1.83 | 0.30 |

| **Characteristic** | **OR<sup>1</sup>** | **95% CI<sup>1</sup>** | **p-value** |
| --- | --- | --- | --- |
| Invasive lobular | 0.90 | 0.11, 8.37 | 0.92 |
| Other/mixed | 0.15 | 0.01, 1.31 | 0.12 |
| **Stage** |  |  |  |
| 0 | — | — |  |
| I | 0.75 | 0.19, 2.82 | 0.67 |
| II | 0.62 | 0.12, 3.06 | 0.56 |
| III | 0.00 |  | 0.99 |
| **pT** |  |  |  |
| is | — | — |  |
| ypT0 | 0.00 |  | 0.99 |
| 1 | 0.75 | 0.19, 2.82 | 0.67 |
| 2 | 0.60 | 0.10, 3.24 | 0.55 |
| 3 | 0.75 | 0.03, 21.6 | 0.85 |
| **pN** |  |  |  |
| 0 | — | — |  |
| 1 | 2.09 | 0.19, 46.7 | 0.56 |
| **Size (cm)** | 1.03 | 0.67, 1.62 | 0.87 |
| **Grade** | 0.72 | 0.34, 1.47 | 0.37 |
| **Close or positive margin** | 2.19 | 0.39, 17.0 | 0.39 |
| **Positive lymph nodes (no)** | 2.07 | 0.53, 34.9 | 0.41 |
| **Lymph nodes removed (no)** | 0.96 | 0.69, 1.33 | 0.81 |
| **ER positive** | 4.38 | 0.59, 89.4 | 0.20 |
| **PR positive** | 0.95 | 0.20, 4.53 | 0.95 |
| **HER2 status** |  |  |  |
| Positive | — | — |  |
| Negative | 0.94 | 0.10, 8.64 | 0.95 |
| Not assessed | 1.00 | 0.10, 10.4 | 1.0 |
| Triple negative | 0.32 | 0.02, 2.70 | 0.34 |

<sup>1</sup>OR = Odds Ratio, CI = Confidence Interval

**Supplementary Table 2.** Univariate analysis of treatment characteristics and use of rehabilitation services.

| Characteristic | OR<sup>1</sup> | 95% CI<sup>1</sup> | p-value |
| --- | --- | --- | --- |
| **Axillary Surgery** |  |  |  |
| None | — | — |  |
| Sentinel node | 1.00 | 0.29, 3.49 | 1.0 |
| **Neoadjuvant chemo** | 0.00 |  | 0.99 |
| **Endocrine therapy** |  |  |  |
| Tamoxifen | — | — |  |
| Aromatase Inhibitor | 0.41 | 0.07, 1.83 | 0.26 |
| None | 0.17 | 0.03, 0.86 | 0.04 |
| **Adjuvant chemo** | 1.00 | 0.21, 4.74 | 1.0 |
| **Breast dose (Gy)** | 1.02 | 0.86, 1.20 | 0.84 |
| **Boost dose (Gy)** | 0.99 | 0.88, 1.11 | 0.91 |
| **Fractionation** |  |  |  |
| Conventionally fractionated | — | — |  |
| Hypofractionated | 0.85 | 0.28, 2.60 | 0.78 |
| **Duration of radiation (days)** | 1.01 | 0.95, 1.07 | 0.84 |
| **Radiation Fields** |  |  |  |
| Tangents | — | — |  |
| High Tangents | 1.00 | 0.27, 3.77 | 1.0 |
| Regional nodal irradiation | 1.00 | 0.04, 26.6 | 1.0 |

<sup>1</sup>OR = Odds Ratio, CI = Confidence Interval

## References

1. Street W. Breast Cancer Facts & Figures 2019–2020.

2. Cancer Treatment & Survivorship Facts & Figures 2022–2024.

3. Hopwood P, Haviland JS, Sumo G, et al. Comparison of patient-reported breast, arm, and shoulder symptoms and body image after radiotherapy for early breast cancer: 5-year followup in the randomised Standardisation of Breast Radiotherapy (START) trials. Lancet Oncol. 2010;11(3):231–240. doi:10.1016/S1470-2045(09)70382-1

4. Rietman JS, Dijkstra PU, Hoekstra HJ, et al. Late morbidity after treatment of breast cancer in relation to daily activities and quality of life: a systematic review. Eur J Surg Oncol J Eur Soc Surg Oncol Br Assoc Surg Oncol. 2003;29(3):229–238. doi:10.1053/ejso.2002.1403

5. Nesvold IL, Fosså SD, Holm I, Naume B, Dahl AA. Arm/shoulder problems in breast cancer survivors are associated with reduced health and poorer physical quality of life. Acta Oncol. 2010;49(3):347–353. doi:10.3109/02841860903302905

6. Tengrup I, Tennvall-Nittby L, Christiansson I, Laurin M. Arm morbidity after breast-conserving therapy for breast cancer. Acta Oncol Stockh Swed. 2000;39(3):393–397. doi:10.1080/028418600750013177

7. Boquiren VM, Hack TF, Thomas RL, et al. A longitudinal analysis of chronic arm morbidity following breast cancer surgery. Breast Cancer Res Treat. 2016;157(3):413–425. doi:10.1007/s10549-016-3834-8

8. Lauridsen MC, Christiansen P, Hessov I. The effect of physiotherapy on shoulder function in patients surgically treated for breast cancer: a randomized study. Acta Oncol Stockh Swed. 2005;44(5):449–457. doi:10.1080/02841860510029905

9. Lipps DB, Sachdev S, Strauss JB. Quantifying radiation dose delivered to individual shoulder muscles during breast radiotherapy. Radiother Oncol. 2017;122(3):431–436. doi:10.1016/j.radonc.2016.12.032

10. Wallner C, Drysch M, Hahn SA, et al. Alterations in pectoralis muscle cell characteristics after radiation of the human breast in situ. J Radiat Res (Tokyo). 2019;60(6):825–830. doi:10.1093/jrr/rrz067

11. Lipps DB, Leonardis JM, Dess RT, et al. Mechanical properties of the shoulder and pectoralis major in breast cancer patients undergoing breast-conserving surgery with axillary surgery and radiotherapy. Sci Rep. 2019;9(1):17737. doi:10.1038/s41598-019-54100-6

12. Wolfram S, Takayesu JSK, Pierce LJ, Jagsi R, Lipps DB. Changes in pectoralis major stiffness and thickness following radiotherapy for breast cancer: A 12-month follow-up case series. Radiother Oncol J Eur Soc Ther Radiol Oncol. 2022;179:109450. doi:10.1016/j.radonc.2022.109450

13. Seo A, Hwang JM, Lee JM, Jung TD. Changes in Pectoral Muscle Volume During Subacute Period after Radiation Therapy for Breast Cancer: A Retrospective up to 4-year Follow-up Study. Sci Rep. 2019;9(1):7038. doi:10.1038/s41598-019-43163-0

14. Shamley DR, Srinanaganathan R, Weatherall R, et al. Changes in shoulder muscle size and activity following treatment for breast cancer. Breast Cancer Res Treat. 2007;106(1):19–27. doi:10.1007/s10549-006-9466-7

15. Yang EJ, Park WB, Seo KS, Kim SW, Heo CY, Lim JY. Longitudinal change of treatment-related upper limb dysfunction and its impact on late dysfunction in breast cancer survivors: A prospective cohort study. J Surg Oncol. 2010;101(1):84–91. doi:10.1002/jso.21435

16. Hughes KS, Schnaper LA, Berry D, et al. Lumpectomy plus Tamoxifen with or without Irradiation in Women 70 Years of Age or Older with Early Breast Cancer. N Engl J Med. 2004;351(10):971–977. doi:10.1056/NEJMoa040587

17. Matzinger O, Heimsoth I, Poortmans P, et al. Toxicity at three years with and without irradiation of the internal mammary and medial supraclavicular lymph node chain in stage I to III breast cancer (EORTC trial 22922/10925). Acta Oncol. 2010;49(1):24–34. doi:10.3109/02841860903352959

18. Poortmans PM, Collette S, Kirkove C, et al. Internal Mammary and Medial Supraclavicular Irradiation in Breast Cancer. N Engl J Med. 2015;373(4):317–327. doi:10.1056/NEJMoa1415369

19. Whelan TJ, Olivotto IA, Parulekar WR, et al. Regional Nodal Irradiation in Early-Stage Breast Cancer. N Engl J Med. 2015;373(4):307–316. doi:10.1056/NEJMoa1415340

20. Bruce J, Mazuquin B, Canaway A, et al. Exercise versus usual care after non-reconstructive breast cancer surgery (UK PROSPER): multicentre randomised controlled trial and economic evaluation. BMJ. 2021;375:e066542. doi:10.1136/bmj-2021-066542

21. Paolucci T, Bernetti A, Bai AV, et al. The sequelae of mastectomy and quadrantectomy with respect to the reaching movement in breast cancer survivors: evidence for an integrated rehabilitation protocol during oncological care. Support Care Cancer Off J Multinatl Assoc Support Care Cancer. 2021;29(2):899–908. doi:10.1007/s00520-020-05567-x

22. Wiskemann J, Schmidt ME, Klassen O, et al. Effects of 12-week resistance training during radiotherapy in breast cancer patients. Scand J Med Sci Sports. 2017;27(11):1500–1510. doi:10.1111/sms.12777

23. Lim SM, Han Y, Kim SI, Park HS. Utilization of bioelectrical impedance analysis for detection of lymphedema in breast Cancer survivors: a prospective cross sectional study. BMC Cancer. 2019;19(1):669. doi:10.1186/s12885-019-5840-9

24. Glassman GE, Dellalana L, Tkaczyk ER, et al. Measuring Biomechanical Properties Using a Noninvasive Myoton Device for Lymphedema Detection and Tracking: A Pilot Study. Eplasty. 2022;22:e54.

